# Disparities in US public historic cancer mortality data: Advocacy for gastrointestinal cancers, tailored prevention measures, and the inclusion of oversea deaths. -Under-reporting of cancer deaths as sign of health disparity

**DOI:** 10.1101/2025.01.22.25320955

**Authors:** Tingting Zhang, Juan Valle, Ashokkumar A. Patel, Stacie Lindsey, Aki Smith, Tony Kin Wai Hung

**Affiliations:** Hear2Care; Colangiocarcinoma Foundation; Hope for Stomach Cancer

## Abstract

Historically, data from US death certificates, available through the CDC’s WONDER database, have been used to highlight health disparities. The 5 year (2018-2022) underlying-cause-of-death data were analyzed for different race/ethnicity groups. Gastrointestinal (GI) cancers of the colon/rectum, pancreas, liver/bile duct, and stomach contributed to more than a quarter of cancer deaths in the US, calling for focused advocacy. Known disparities for Black non-Hispanic Americans were verified in cancers of the colon/rectum, and pancreas. However, mortality data for “More than one race” non-Hispanic group or non-White Hispanic group appeared unreliable, suggesting that under-reporting is also a sign of health disparity. Age-specific death rates (ASDRs) were calculated to view health disparities in various age groups. Cancers of the colon/rectum, liver, and stomach cause significant mortality in the under-50 population. And minority groups are more likely to die from cancers in the liver or the stomach compared to the White non-Hispanics. Liver cancer crude death rate was lower than expect in Asian Non-Hipanics when compared with the high mortalities of IARC’s GLOBOCAN estimates for Asian countries. Census data were then used to calculate Asian sub-groups’ ASDRs. Significantly higher risks were seen in gastric cancer for Korean Americans and liver cancer for Vietnamese Americans. The Asian Indian group had the lowest death rates across several GI cancers, even in gallbladder cancer. Some immigrants go back to their birth countries at end of life and these deaths are not reflected in WONDER because consulate reports of American citizens’ deaths abroad do not include race/ethnicity data.

**SIGNIFICANCE:** Gastrointestinal cancers should be advocacy focuses to promote cancer equity. Under-reporting of deaths for minority groups is a sign of health disparity. ASDRs (Age Specific Death Rates) provide bases for tailored screening and prevention. Exclusion of American citizen’s deaths abroad in U.S. public health datasets may mask care access issues faced by immigrant cancer patients who may die overseas. Updating how these deaths are reported will help address this issue.

## INTRODUCTION

Patient advocacy groups in the US need accurate patient outcomes data to focus advocacy efforts. However, many advocacy groups may lack expertise in accessing available epidemiology data, exemplified by a question raised in the audience during the September 8th 2023 NCI Cancer Health Equity Visioning Minilab: “Can we have a bird!s-eye view of health disparity in cancer care in the US?”.

Historically, cancer health disparities have been assessed using data from cancer registries, such as those from the Surveillance, Epidemiology, and End Results (SEER) [ref Ries LG] and the U.S. Cancer Statistics (USCS) where SEER data was combined with those from Center of Disease Control (CDC)’s National Program of Cancer Registries (NPCR). A patient advocate can access the USCS Data Visualizations tool to visualize both age-adjusted incidents (new cancer) rates or age-adjusted death rates (AADR) for each race/ethnic groups such as Hispanic, or non-Hispanic single race groups - White, Black, American Indian and Alaskan Native (AI/AN), Asian, Native Hawaiian and Other Pacific Islander (NH/OPI). This top-level view, adjusting age-specific rates to a “standard” population may not provide in-depth views on the sub-groups the advocacy groups are interested in.

Since survival/death is the most frequently used outcome measure for cancer, cancer patient advocates can also look into the annual estimated cancer deaths published by American Cancer Society [ref Islami F] or the annual report on deaths by the National Vital Statistics Systems [ref Kochanek KD]. Reports of cancer deaths in different race/ethnic groups can be obtained from the Wide-ranging ONline Data for Epidemiologic Research (WONDER) system from CDC[ref Friede A]. With the adoption of the 2003 version of death certificates by all the states in the US from 2018, we now have a “birds-eye” view of health disparities in the past 5 years (2018-2022) from the underlying-cause of death data in different cancers, different age groups, and different race/ethnicities.

In this article, we aims to address three questions related to health disparities in cancer care: (1) what do historical death data reveal about gaps in our national priorities on addressing health disparities? (2) what challenges are present in the quality of data collection, particularly in immigrant populations?; (3) how can we improve data quality to better inform advocacy efforts.

## METHODS

### US Cancer Statistics

We downloaded top ten cancer types and associated AADRs 2017-2021 for both sexes in table formats from the USCS Data Visualizations tool (https://gis.cdc.gov/Cancer/USCS/#/AtAGlance/) for all available race/ethnic groups to take an initial view of top cancer types and assess potential data quality issues.

### WONDER @ CDC

We queried the “Underlying Cause of Death, 2018-2022, Single Race” dataset for “CD-10 Codes: C00-D48 (Neoplasms)” in WONDER@CDC (https://wonder.cdc.gov/ucd-icd10-expanded.html) [CDC WONDER (RRID:SCR_025830)] with the following parameter combinations:

“Group By: Cause of death” (for total), or “Group By: Cause of death; Single Race 6; Hispanic Origin” with “Rate per 100,000” based on “Default intercensal populations for years 2001-2009 (except Infant Age Groups)” to calculate total numbers of death and associated crude death rates from 2018 to 2022;

“Group By: Cause of death; Single Race 15; Hispanic Origin” with “Single Race 15: Asian Indian; Chinese; Filipino; Japanese; Korean; Vietnamese; Other Asian” to calculate deaths from GI cancers for Non-Hispanic Asian groups (population data were not available);

“Group By: Cause of death; Five-Year Age Groups”, or “Group By: Cause of death; Five-Year Age Groups; Single Race 6; Hispanic Origin” in “Five-Year Age Groups: 1-4; 5-9; 10-14; 15-19; 20-24; 25-29; 30-34; 35-39; 40-44; 45-49; 50-54; 55-59; 60-64; 65-69; 70-74; 75-79; 80-84; 85-89; 90-94; 95-99” based on “Default intercensal populations for years 2001-2009 (except Infant Age Groups)” to calculate death numbers and age-specific death rates in 5 year age-groups;

“Group By: Cause of death; 10-Year Age Groups; Single Race 15; Hispanic Origin” in “10-Year Age Groups: 1-4; 5-14; 15-24; 25-34; 35-44; 45-54; 55-64; 65-74 ; 75--84; 85+with “Single Race 15: Asian Indian; Chinese; Filipino; Japanese; Korean; Vietnamese; Other Asian” to calculate death numbers from GI cancers in 10-year age groups.

The death numbers for GI cancers were calculated with deaths of these ICD10 codes [International Classification of Diseases Version 10 (RRID:SCR_010349)] combined:

Esophageal Cancer: C15.0, C15.1, C15.2, C15.3, C15.4, C15.5, C15.8, C15.9;

Stomach Cancer: C16.0, C16.1, C16.2, C16.3, C16.4, C16.5, C16.6, C16.8, C16.9;

Cancer in Rectum and Colon (CRC): C18.0, C18.2, C18.3, C18.4, C18.5, C18.6, C18.7, C18.8, C18.9, C19. C20;

Cancer in Liver and Intrahepatic Bile Duct: C22;

Liver Cancer: C22.0, C22.2, C22.3, C22.4, C22.9;

Bile Duct Cancer: C22.1, C24.0, C24.,8, C24.9;

Gallbladder Cancer: C23;

Pancreatic Cancer: C25.0, C25.1, C25.2, C25.3, C25.4, C25.7, C25.8, C25.9.

It is worth noting that reporting of any death number equal or below 9 is suppressed to protect privacy therefore death numbers combining several sub-categories may under-report real death numbers. The 10-year groups were chosen for the smaller Asian groups to minimize the impact of data suppression.

### US Census

Because no population data was provided by WONDER@CDC for the “Single Race 15” Asian groups, the denominators to calculate death rates were created by combining total population data from ACS S0201 table (https://data.census.gov/table/ACSSPP1Y2022.S0201) for years 2018, 2019, 2021 and 2022, and the Decennial Census table P1 (https://data.census.gov/table/DECENNIALCD1182020.P1) for 2020. Age-specific population data for these Asian groups to calculate ASDR were obtained from the “DP05:ACS 2021 5 years estimates selected population table”(https://data.census.gov/table?q=DP05) and merged to form the 15-24 and 55-64 10-year age groups. [U.S. Census Bureau (RRID:SCR_011587)].

### GLOBOCAN

Estimated crude death rates in 2022 for both sexes for cancers in the liver and intrahepatic bile duct (ICD10:C22) in countries where many of Asian Americans trace heritages to were downloaded from GLOBOCAN CANCER TODAY DATAVIZ tool (https://gco.iarc.fr/today/en/dataviz/tables?mode=population&types=1&cancers=11). [Global Cancer Observatory Cancer Today (RRID:SCR_025452)] Caution must be exercised since the mortality data quality varies. For Japan, South Korea, Singapore and Mongolia, the rates were those from 2010-2019 projected to 2022 and applied to 2022 population. For the rest, the rates were estimated from incidence (weighted average of either sub-national or urban registries applied to 2022 population) using mortality:incidence ratios derived from survival estimation, thus with lower quality.

## RESULTS

### Top ten cancer types from USCS

Gastrointestinal cancers contribute to 3 (in White or Black non-Hispanic(WNH, BNH) population) to 4 (in Hispanic or Asian, AI/AN, HA/OPI non-Hispanic) of the 10 cancer types with the highest mortality (AADR) in the US (Table 1). AADRs of BNH population are the highest in all populations for cancers in male prostate, female breast, colon and rectum, pancreas, and uterus. Assignment of correct race groups on US death certificates were reported to be excellent for WNH or BNH populations, and good (3% off) for Hispanic, or ANH population. Therefore, the lower AADR across the board for the latter two groups, which contains a large percentage of immigrants, might be the sign of a different reporting issue from those of AI/AN non-Hispanic population whose race assignment can be off by 30% on death certificates [ref Arias E]. Despite being high on this death list, GI cancers especially cancers in the liver and bile duct, or stomach, are currently amongst the least funded by the nation’s federal or private research grants [ref Haghighat S].

**Table 1.** Top ten lists of cancer types that caused highest mortality in the form of age-adjusted death rate (AADR) from US Cancer Statistics for White non-Hispanic, Black non-Hispanic, Hispanic, Asian non-Hispanic, American Indian and Alaska Native (AI/AN) non-Hispanic, and Native Hawaiian and Other Pacific Islander (NH/OPI) non-Hispanic groups.

| USCS TopTen 2018-2022 Both Sexes |  |  |  |  |  |  |  |  |  |  |  |
| --- | --- | --- | --- | --- | --- | --- | --- | --- | --- | --- | --- |
| White Non-Hispanic |  | Black Non-Hispanic |  | Hispanic |  | Asian Non-Hispanic |  | AIAN Non-Hispanic |  | NHOPi Non-Hispanic |  |
| Cancer Type | AADR | Cancer Type | AADR | Cancer Type | AADR | Cancer Type | AADR | Cancer Type | AADR | Cancer Type | AADR |
| Lung and Bronchus | 35.4 | Prostate | 37.3 | Prostate | 15.2 | Lung and Bronchus | 18.4 | Lung and Bronchus | 27.3 | Lung and Bronchus | 27.1 |
| Female Breast | 19.4 | Lung and Bronchus | 34.6 | Lung and Bronchus | 14.5 | Female Breast | 11.4 | Female Breast | 14.7 | Female Breast | 24.3 |
| Prostate | 18 | Female Breast | 27.1 | Female Breast | 13.6 | **Colon and Rectum** | 9 | Prostate | 13.4 | Prostate | 16.3 |
| **Colon and Rectum** | 12.9 | **Colon and Rectum** | 16.9 | **Colon and Rectum** | 10.6 | Prostate | 8.3 | **Colon and Rectum** | 12.9 | Corpus and Uterus NOS | 14.4 |
| **Pancreas** | 11.4 | **Pancreas** | 13.8 | **Liver and Intrahepatic Bile Duct** | 9 | **Liver and Intrahepatic Bile Duct** | 8 | **Liver and Intrahepatic Bile Duct** | 9.4 | **Colon and Rectum** | 11.6 |
| Ovary | 6.3 | Corpus and Uterus NOS | 9.6 | **Pancreas** | 8.9 | **Pancreas** | 7.6 | **Pancreas** | 8.5 | **Liver and Intrahepatic Bile Duct** | 10.5 |
| Leukemias | 6.3 | **Liver and Intrahepatic Bile Duct** | 8 | Ovary | 4.8 | Ovary | 4.3 | Kidney and Renal Pelvis | 4.8 | **Pancreas** | 9.8 |
| **Liver and Intrahepatic Bile Duct** | 5.9 | Myeloma | 5.8 | **Stomach** | 4.6 | **Stomach** | 4.2 | Ovary | 4.8 | Ovary | 5.7 |
| Non-Hodgkin Lymphoma | 5.2 | Ovary | 5.5 | Corpus and Uterus NOS | 4.4 | Non-Hodgkin Lymphoma | 3.5 | Corpus and Uterus NOS | 3.9 | Leukemias | 5.7 |
| Brain and Other Nervous System | 5.1 | Leukemias | 5.1 | Non-Hodgkin Lymphoma | 4.3 | Corpus and Uterus NOS | 3.4 | **Stomach** | 3.9 | **Stomach** | 5.5 |

### Overview of Disparity and Under-Reporting in GI Cancers from WONDER@CDC

Underlying cause of death numbers of GI cancers from 2018-2022 for different race/ethnic groups were collected from WONDER@CDC and crude death rates were calculated against reference populations data (Table 2). It is clear that crude death rate (CDR) for neither “non-White Hispanic” group nor “More than one race non-Hispanic” group is reliable. CDRs are different from USCS’s AADRs because the latter was adjusted according to the different age distribution of these groups. Each of these GI cancers causes five-figure deaths a year on average (close to 10,000 for bile duct cancer; over 10,000 for stomach cancer; over 15,000 for oesophageal cancer; 20,000 for liver cancer; and around 50,000 for either CRC or pancreatic cancer) and contribute to more than a quarter of total annual cancer deaths.

**Table 2.** Comparison of the numbers of deaths and crude death rate (CDR) from WONDER@CDC for 6 major gastrointestinal (GI) cancers in esophagus, stomach, colon and rectum (CRC), pancreas, liver, and bile duct (CCA) for American Indian and Alaska Native (AI/AN) non-Hispanic, Asian non-Hispanic, Black non-Hispanic, Hispanic, More than one race non-Hispanic, Native Hawaiian and Other Pacific Islander (NH/OPI) non-Hispanic, White non-Hispanic, White Hispanic, non-White Hispanic groups.

|  | Population | Oesophagus |  | Stomach |  | CRC |  | Pancreas |  | Liver |  | Bile Duct |  |
| --- | --- | --- | --- | --- | --- | --- | --- | --- | --- | --- | --- | --- | --- |
|  | 2018-2022<br>5 Years | Deaths | CDR | Deaths | CDR | Deaths | CDR | Deaths | CDR | Deaths | CDR | Deaths | CDR |
| American Indian and Alaska Native Non-Hispanic | 12157505 | 375 | 3.08 | 460 | 3.78 | 1524 | 12.54 | 1052 | 8.65 | 998 | 8.21 | 302 | 2.48 |
| Asian Non-Hispanic | 96963677 | 1564 | 1.61 | 4225 | 4.36 | 9063 | 9.35 | 7628 | 7.87 | 5865 | 6.05 | 2784 | 2.87 |
| Black or African American Non-Hispanic | 207406059 | 6030 | 2.91 | 9528 | 4.59 | 34309 | 16.54 | 28640 | 13.81 | 13803 | 6.66 | 4939 | 2.38 |
| Native Hawaiian or Other Pacific Islander Non-Hispanic | 3057935 | 62 | 2.03 | 142 | 4.64 | 306 | 10.01 | 257 | 8.40 | 215 | 7.03 | 83 | 2.71 |
| More than one race Non-Hispanic | 37729938 | 279 | 0.74 | 289 | 0.77 | 1136 | 3.01 | 886 | 2.35 | 549 | 1.46 | 223 | 0.59 |
| White Non-Hispanic | 984689016 | 66305 | 6.73 | 30409 | 3.09 | 188082 | 19.10 | 176811 | 17.96 | 64503 | 6.55 | 35293 | 3.58 |
| White Hispanic or Latino | 269345779 | 3858 | 1.43 | 9668 | 3.59 | 19791 | 7.35 | 17515 | 6.50 | 14422 | 5.35 | 5177 | 1.92 |
| Non-White Hispanic | 38722473 | 115 | 0.30 | 237 | 0.61 | 647 | 1.67 | 577 | 1.49 | 398 | 1.03 | 148 | 0.38 |
| **Total** | **1650072382** | **78813** | **4.78** | **55115** | **3.34** | **255377** | **15.48** | **233804** | **14.17** | **101103** | **6.13** | **49085** | **2.97** |

By looking into age-specific death rate (ASDR), calculating deaths in 100,000 for each age groups against the relevant populations, we have a more detailed view of cancer risks across the age groups for the different race/ethnic groups. The ASDRs in a smaller population such at the NH/OPI NH group was impacted significantly by data suppression and thus not shown.

WNH population appeared to be more susceptible to deaths caused by oesophageal cancer across all age groups than any other race/ethnic population (Figure 1 A). On the other hand, all minority race/ethnic groups have higher ASDRs in stomach cancer than WNH group (Figure 1B), highlighting a contrasted disparity for cancers in the upper digestive track.

**Figure 1.**
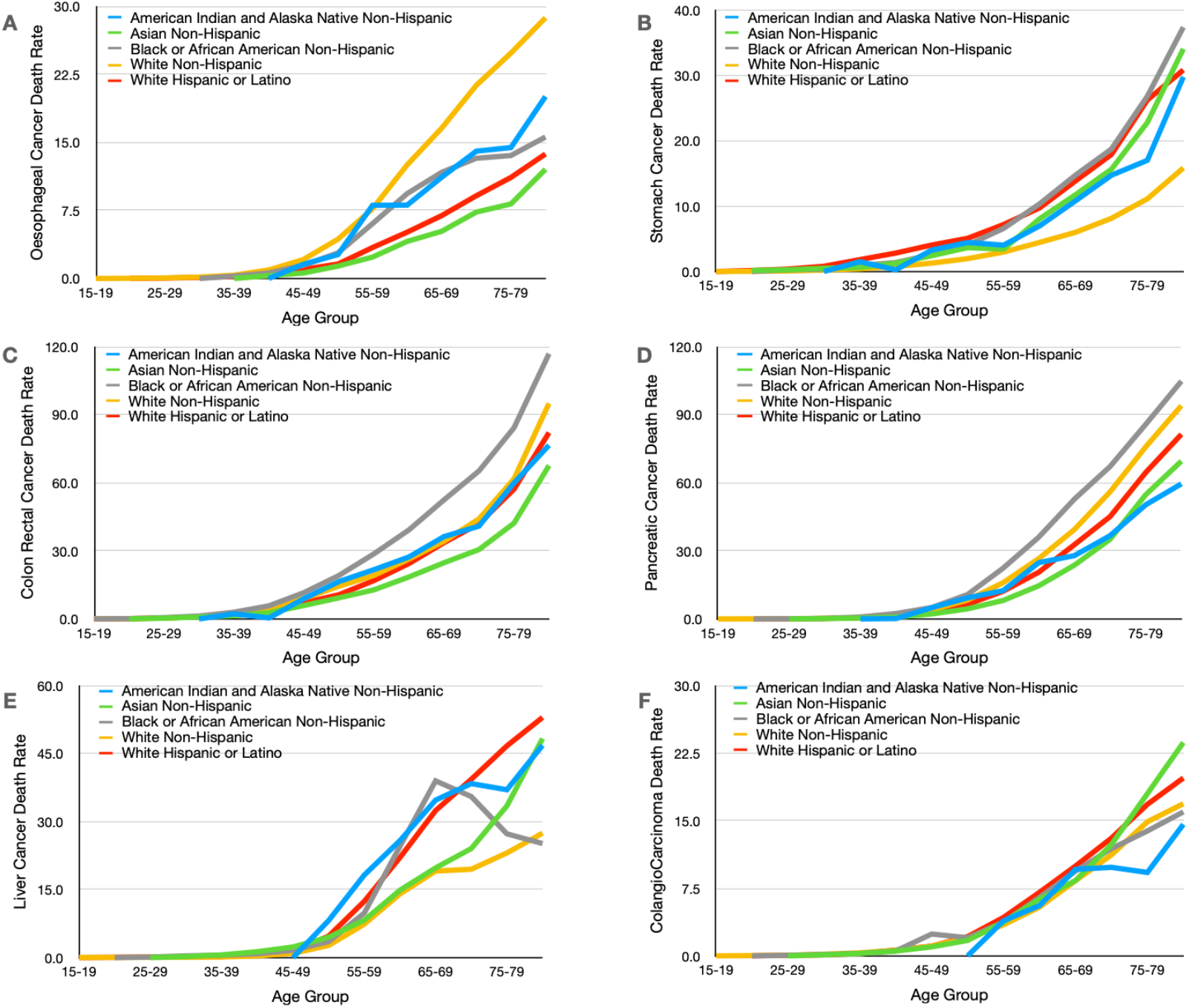
Visualisation of age-specific death rates (ASDRs) in 5-year age groups from WONDER@CDC in a line graph for 6 major gastrointestinal (GI) cancers in A: esophagus, B: stomach, C: colon and rectum (CRC), D: pancreas, E: liver, and F: bile duct (CCA) for American Indian and Alaska Native (AI/AN) non-Hispanic, Asian non-Hispanic, Black non-Hispanic, Hispanic, Native Hawaiian and Other Pacific Islander (NH/OPI) non-Hispanic, White non-Hispanic, White Hispanic groups.

Validating what we see from AADR in USCS, the BNH group have the highest ASDR of colon and rectum cancers (CRC) and pancreatic cancer (Figure 1C, 1D). ASDR of CRC for WNH, WH, and AI/AN NH appeared to be similar, while ASDR of CRC for ANH trended lower. ASDRs for CRC in the overall under-50 age population are already quite high: 4.5/100,000 for 40-44 and 8.6/100,000 for 45-49 age groups in total population (Supplement 1), calling for earlier screening and prevention measures.

Since cancers in the liver and cancers in bile ducts have different risk factors, their ASDRs were analyzed separately. ASDRs of liver cancer for WH, AI/AN NH groups are consistently high than those for BNH, The most surprising ASDRs of liver cancer were for the ANH group, which despited being high until 55, flattened out to be close to those of WNH group before trending high again ((Figure 1E). This abnormality, together with the observation of lower than expected AADR in USCS (Table 1), motivated us to look into the ASDRs of “Single Race 15” disaggregated Asian groups. ASDRs of bile duct cancer (CCA) for ANH, WH groups appeared higher from other race/ethnic groups, especially in the older age-groups (Figure 1F).

It is also worth noting that despite having lower ASDR, 3 out of 5 deaths in either stomach cancer or liver cancer were in the WNH group (Table 2). CRC, stomach, bile duct, and liver cancers each take hundreds of lives under-40 annually, and it goes up to thousands for stomach cancer and CRC for those under 50 (Supplement 1).

**Supplement 1.** Five years total of age-specific numbers of deaths and death rates for the US population 2018-2022 from WONDER@CDC for f6 major gastrointestinal (GI) cancers in esophagus, stomach, colon and rectum (CRC), pancreas, liver, and bile duct (CCA).

| Age Group | <1 | 1-4 | 5-9 | 10-14 | 15-19 | 20-24 | 25-29 | 30-34 | 35-39 | 40-44 | 45-49 | 50-54 | 55-59 | 60-64 | 65-69 | 70-74 | 75-79 | 80-84 |
| --- | --- | --- | --- | --- | --- | --- | --- | --- | --- | --- | --- | --- | --- | --- | --- | --- | --- | --- |
| **Oesophageal** |  |  |  |  |  | 19 | 55 | 138 | 321 | 777 | 1670 | 3561 | 7098 | 11257 | 12912 | 13542 | 10844 | 8103 |
| **Stomach** |  |  |  |  | 15 | 70 | 205 | 418 | 829 | 1313 | 2019 | 2903 | 4381 | 6134 | 7142 | 7497 | 7080 | 6325 |
| **Colon/Rectum** |  |  |  |  | 32 | 134 | 468 | 1192 | 2618 | 4577 | 8630 | 14419 | 21019 | 28007 | 31428 | 32437 | 30625 | 29708 |
| **Pancreas** |  |  |  |  | 12 | 20 | 60 | 227 | 666 | 1668 | 3815 | 8276 | 16900 | 27254 | 34920 | 39638 | 36809 | 29289 |
| **Liver** | 12 | 57 | 33 | 15 | 52 | 64 | 112 | 179 | 292 | 552 | 1227 | 3343 | 9039 | 16764 | 20082 | 16564 | 12744 | 9594 |
| **Bile Duct** |  |  |  |  |  | 30 | 70 | 194 | 332 | 641 | 1069 | 2050 | 3853 | 5928 | 7642 | 8261 | 7431 | 5470 |

**B** Age-Specific Death Rate, GI Cancers, 2018-2022
| Age Group | <1 | 1-4 | 5-9 | 10-14 | 15-19 | 20-24 | 25-29 | 30-34 | 35-39 | 40-44 | 45-49 | 50-54 | 55-59 | 60-64 | 65-69 | 70-74 | 75-79 | 80-84 |
| --- | --- | --- | --- | --- | --- | --- | --- | --- | --- | --- | --- | --- | --- | --- | --- | --- | --- | --- |
| **Oesophageal** |  |  |  |  |  | 0.02 | 0.05 | 0.12 | 0.29 | 0.76 | 1.66 | 3.44 | 6.57 | 10.82 | 14.44 | 18.67 | 21.83 | 25.43 |
| **Stomach** |  |  |  |  | 0.01 | 0.06 | 0.18 | 0.37 | 0.76 | 1.28 | 2.01 | 2.81 | 4.06 | 5.89 | 7.99 | 10.34 | 14.25 | 19.85 |
| **Colon/Rectum** |  |  |  |  | 0.03 | 0.12 | 0.41 | 1.05 | 2.39 | **4.47** | **8.59** | 13.94 | 19.47 | 26.91 | 35.14 | 44.72 | 61.65 | 93.22 |
| **Pancreas** |  |  |  |  | 0.01 | 0.02 | 0.05 | 0.20 | 0.61 | 1.63 | 3.80 | 8.00 | 15.65 | 26.19 | 39.04 | 54.64 | 74.10 | 91.90 |
| **Liver** | 0.06 | 0.07 | 0.03 | 0.01 | 0.05 | 0.06 | 0.10 | 0.16 | 0.27 | 0.54 | 1.22 | 3.23 | 8.37 | 16.11 | 22.45 | 22.83 | 25.66 | 30.10 |
| **Bile Duct** |  |  |  |  |  | 0.03 | 0.06 | 0.17 | 0.30 | 0.63 | 1.06 | 1.98 | 3.57 | 5.70 | 8.54 | 11.39 | 14.96 | 17.16 |

### The Need for Disaggregation of the Asian Data and Signs of Data-Missing

The Asian non-Hispanic population includes a large percentage of immigrants from countries/regions where deaths from liver cancer were amongst the highest in the world based on GLOBOCAN [ref Bray E] (Supplement 2). Vietnam, China, South Korea, and the Philippines have high estimate death rates, and especially higher are those of Mongolia, Thailand, Cambodia, and Laos. We went on to investigate the lower than expected liver cancer mortality in this group (Table1) with disaggregated Asian American 5-years total death numbers (2018-2022) available in the “Single Race 15” group from WONDER@CDC for aforementioned six GI cancers as well as gallbladder cancer which is a known risk for Asian Indians (Table 3). Population data from 2018-2022 for each Asian alone groups were collected from US Census tables as the denominators to calculate crude death rates (CDRs). The Japanese group had the highest CDRs and Asian Indians had the lowest CDRs across almost all GI cancer types, even for bladder cancer. Based on US Census DP05 ACS 2021 estimates (Supplement 3), these two groups have distinctly different age-distributions. The median age for the Japanese group is 52.3 yo and that of the Asian Indian groups is 34.5, where as the media ages for other Asian groups fall between 38-43, We then collected the 5-year total death numbers from 6 GI cancers in 10 year age groups and calculated and plotted ASDRs for 7 Asian groups against those of the WNH group (Figure 2). We are providing a template using liver cancer as an example in the Supplement (Supplement data) so other patient advocates interested in other cancers or other diseases can run the same visualisation.

**Table 3.** Comparison of the numbers of deaths and crude death rate (CDR) for 7 gastrointestinal (GI) cancers in esophagus, stomach, colon and rectum (CRC), pancreas, liver, bile duct (CCA), and gallbladder for Asian Indian, Chinese, Filipino, Japanese, Korean, Vietnamese, and Other Asian groups. *Due to the fact that no relevant population data was provided by WONDER@CDC for these groups, we combined single race alone population data from ACS 2018, 2019, 2021, 2022, and Census 2020 to come up with the denominator to calculate CDR. Other Asian data was a subtraction from total Asian alone data.

|  | Population | Oesophageal Cancer |  | Stomach Cancer |  | Colon Cancer |  | Pancreatic Cancer |  | Liver Cancer |  | Colangio-Carcinoma |  | Gall Bladder Cancer |  |
| --- | --- | --- | --- | --- | --- | --- | --- | --- | --- | --- | --- | --- | --- | --- | --- |
|  | 2018-2022 5 Years (*) | Deaths | Crude Death Rate | Deaths | Crude Death Rate | Deaths | Crude Death Rate | Deaths | Crude Death Rate | Deaths | Crude Death Rate | Deaths | Crude Death Rate | Deaths | Crude Death Rate |
| Asian Indian | 21735853 | 266 | 1.22 | 246 | 1.13 | 719 | 3.31 | 710 | 3.27 | 364 | 1.67 | 323 | 1.49 | 88 | **0.40** |
| Chinese | 21811744 | 386 | 1.77 | 1294 | 5.93 | 2372 | 10.87 | 2082 | 9.55 | 1392 | **6.38** | 716 | 3.28 | 135 | 0.62 |
| Filipino | 14910753 | 193 | 1.29 | 416 | 2.79 | 1688 | 11.32 | 1412 | 9.47 | 839 | 5.63 | 433 | 2.90 | 99 | 0.66 |
| Japanese | 3739954 | 199 | 5.32 | 402 | 10.75 | 890 | 23.80 | 924 | 24.71 | 368 | 9.84 | 203 | 5.43 | 46 | 1.23 |
| Korean | 7385599 | 117 | 1.58 | 800 | 10.83 | 1013 | 13.72 | 952 | 12.89 | 589 | 7.97 | 384 | 5.20 | 81 | 1.10 |
| Vietnamese | 9472088 | 176 | 1.86 | 459 | 4.85 | 1012 | 10.68 | 630 | 6.65 | 1331 | 14.05 | 275 | 2.90 | 61 | 0.64 |
| Other Asian | 17907686 | 211 | 1.18 | 523 | 2.92 | 1247 | 6.96 | 831 | 4.64 | 960 | 5.36 | 450 | 2.51 | 96 | 0.54 |
| **Total** | **96963677** | **1548** | **1.60** | **4140** | **4.27** | **8941** | **9.22** | **7541** | **7.78** | **5843** | **6.03** | **2784** | **2.87** | **606** | **0.62** |

**Figure 2.**
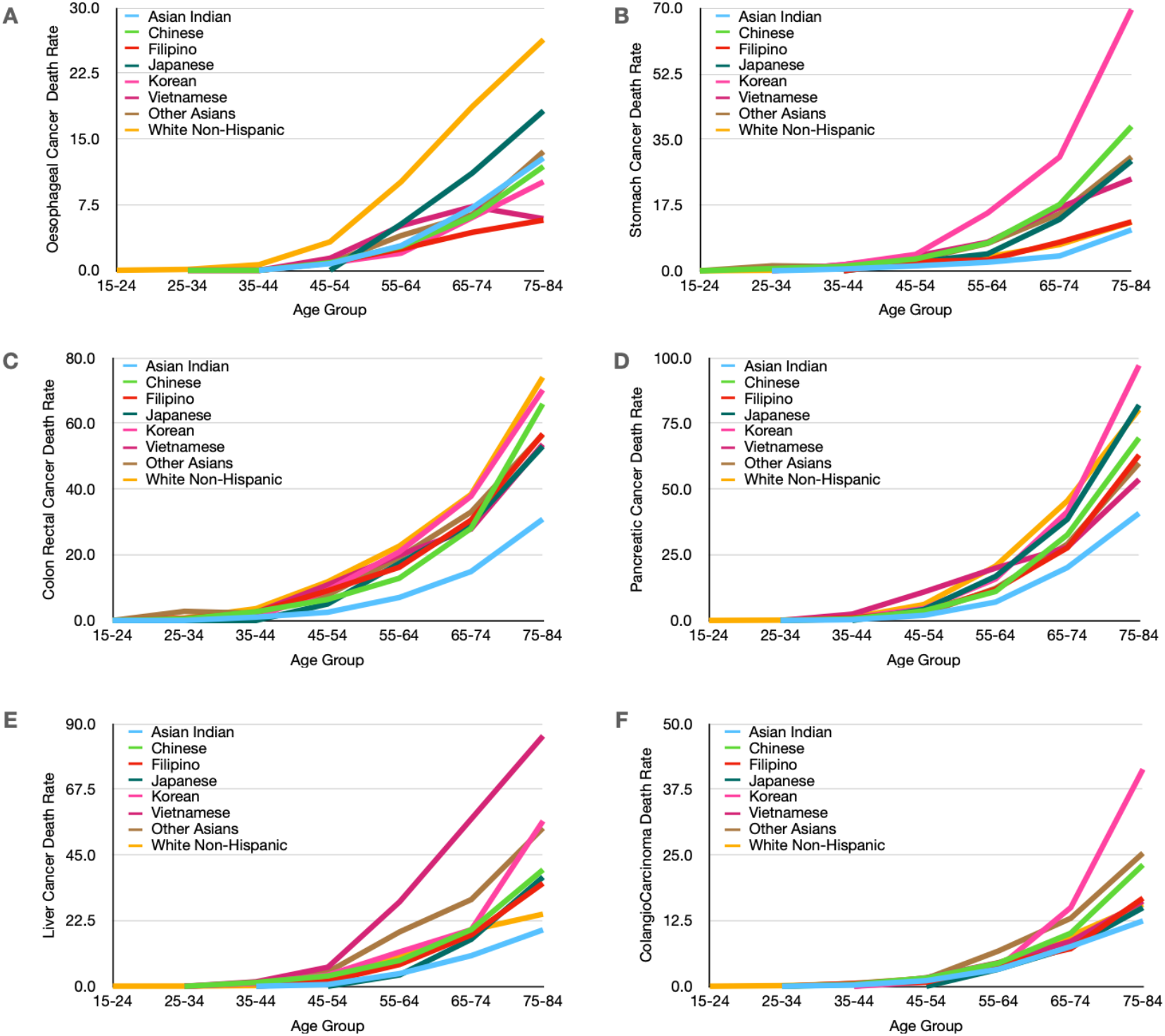
Visualisation of age-specific death rates (ASDRs) in 10-year age groups from WONDER@CDC in a line graph for for 6 major gastrointestinal (GI) cancers in A: esophagus, B: stomach, C: colon and rectum (CRC), D: pancreas, E: liver, and F: bile duct (CCA) for Asian Indian, Chinese, Filipino, Japanese, Korean, Vietnamese, and Other Asian groups. The population data for these age groups was obtained from Census ACS DP05 (Supplement 3)

**Supplement 2.** GLOBOCAN Cancer Today estimate of cancer mortality for cancers in the liver and bile duct for Asian countries where most Asian groups in US claim heritage. *Higher quality data from WHO

| Label | Alpha-3 code | Deaths | Crude death rate | AADR (World) |
| --- | --- | --- | --- | --- |
| China | CHN | 316544 | 22.42 | 12.59 |
| India | IND | 36953 | 2.63 | 2.61 |
| Japan* | JPN | 26420 | 21.04 | 4.3 |
| South Korea* | KOR | 12595 | 24.54 | 10.67 |
| Philippines | PHL | 11653 | 10.36 | 11.59 |
| Viet Nam | VNM | 23333 | 23.58 | 19.25 |
| Singapore* | SGP | 1353 | 22.76 | 11 |
| Thailand | THA | 27143 | 38.73 | 21.99 |
| Laos | LAO | 1313 | 17.55 | 23.65 |
| Cambodia | KHM | 3290 | 19.16 | 23.67 |
| Mongolia* | MNG | 2164 | 64.06 | 80.16 |

**Supplement 3.** DP05 2021 ACS Five Year Estimates Selected Population Profiles from US Census for Asian groups: Other Asian population was calculated by subtracting from Asian non-Hispanic population data with those listed above (using Chinese Excluding Taiwainese data due to the different policies on dual-citizenship)

|  | Population | Male | Female | Sex ratio | Median age | Under 5 | 5 to 9 | 10 to 14 | 15 to 19 | 20 to 24 | 25 to 34 | 35 to 44 | 45 to 54 | 55 to 59 | 60 to 64 | 65 to 74 | 75 to 84 | 85 and over |
| --- | --- | --- | --- | --- | --- | --- | --- | --- | --- | --- | --- | --- | --- | --- | --- | --- | --- | --- |
| Asian NH | 18782924 | 8990767 | 9792157 | 91.8 | 37.5 | 1011688 | 1041715 | 1076829 | 1113591 | 1214621 | 3106802 | 3040011 | 2628854 | 1124006 | 996093 | 1484486 | 674882 | 269346 |
| Chinese Alone | 4403215 | 2067124 | 2336091 | 88.5 | 38.9 | 191856 | 195899 | 218347 | 272301 | 335493 | 733654 | 612213 | 621409 | 303003 | 266684 | 392428 | 174367 | 85561 |
| Chinese Excluding Taiwanese | 4192786 | 1968628 | 2224158 | 88.5 | 38.9 | 182949 | 187197 | 210438 | 263438 | 322839 | 693315 | 572118 | 591697 | 291319 | 255445 | 373396 | 165107 | 83528 |
| Filipino | 2922680 | 1284856 | 1637824 | 78.4 | 42.8 | 95697 | 115442 | 146703 | 162393 | 181931 | 410031 | 453990 | 458579 | 212357 | 196794 | 303911 | 139292 | 45560 |
| Japanese | 760412 | 324011 | 436401 | 74.2 | 52.3 | 21556 | 22992 | 19508 | 22382 | 28682 | 72048 | 105533 | 119479 | 65530 | 62933 | 107863 | 61411 | 50495 |
| Korean | 1469854 | 664667 | 805187 | 82.5 | 41.2 | 54485 | 57929 | 72113 | 82362 | 86641 | 228117 | 243148 | 237815 | 98896 | 94334 | 122555 | 69157 | 22302 |
| Vietnamese | 1862865 | 894087 | 968778 | 92.3 | 40.0 | 77035 | 88196 | 114450 | 130712 | 133590 | 262157 | 256827 | 316856 | 126149 | 109242 | 156420 | 67791 | 23440 |
| Asian Indian Alone | 4278403 | 2233487 | 2044916 | 109.2 | 34.5 | 310123 | 314842 | 273881 | 222341 | 222678 | 840408 | 886095 | 520824 | 177422 | 147844 | 232117 | 104338 | 25490 |
| Other Asian | 3295924 | 1621031 | 1674893 | NA | NA | 269843 | 255117 | 239736 | 229963 | 238260 | 600726 | 522300 | 383604 | 152333 | 129501 | 188224 | 67786 | 18531 |

The WNH group had the highest oesophageal cancer ASDRs over all Asian groups. The Japanese group had higher ASDRs for oesophageal cancer for the over-55 age groups, and the other groups aggregates together. The Korean group had the highest stomach cancer ASDR especially in the over 55 age groups [ref Lee E], followed by the Chinese, the Vietnamese, Other Asians, and the Japanese. ASDR from stomach cancer for the WNH groups aligned with that of the Filipino group, and ASDR for the Asian Indian group was the lowest even though there were already deaths in the 35-44 age group similar to those of Vietnamese or Korean groups.

WNH group also had the highest colon and rectum cancer (CRC) ASDRs against all Asian groups, with Asian groups trending closely across all age groups, except for the Asian Indian group which ASDRs tend to be at least 1/2 of those of other groups. WNH, Korean, and Japanese group also had the higher pancreatic cancer ASDRs, with the Chinese, Filipino, Vietnamese, and Other Asians followed lower. Again, here the Asian Indian group had lower ASDRs ranging from 2/3 to 1/3 of other groups across the age groups.

When liver cancer deaths were disaggregated, the differences for the Asian American groups became strikingly clear. ASDRs for the under 45 age groups were low for all, possibly reflecting the success of HepB vaccination that was approved in the US during 1980s. For those over 45, the Vietnamese group has ASDRs that are 3 times of WNH group’s and followed by the “Other Asians” group that would include Mongolians, Cambodians, Thais and Laotians. Surprisingly, ASDRs of liver cancer for the Chinese and the Korean groups followed closely with those of the WNH group. Again, liver cancer ASDRs for the Asian Indian was the lowest - 2/3 to 1/2 of the WNH group’s. ASDRs of bile duct cancer (Colangiocarcinoma/CCA) was higher amongst “Other Asians” group which include the Thai population that was known to have high risks due to liver fluke infections [ref Yao KJ]. The even higher ASDRs for the Korean group in the above 65 population would need more data to validate. All the other groups appeared to have similar ASDR in most over 45 age groups when there were enough deaths to overcome data suppression.

### Impact of missing data of the Deaths of US Citizens Overseas

It became clear that the low death numbers of several GI cancers for the Asian Indian group, which contributed to 1/5 of the total ANH population, impacted the overall mortality statistics of ANH. Lower than expected ASDRs of liver cancer were also observed for the Chinese group. The two groups are composed of more recent immigrants (both included 70%-foreign-born, and about 40% Asian Indians arrived in the US within the past 10 year). US citizens living abroad were not reported in the Census data, and there was no publicly available data on how many might have died of cancer overseas. Italian researchers found that inclusion of death reports abroad contribute to a 24% increase of AADR in immigrants for all mortalities [Ref Di Napoli A]

Asian countries have booming medical tourism industries [Ref: Fetscherin M, Kim S,] and many have successfully promoted cancer screening and other services world-wide. Survey of cancer patients diagnosed in Kenya’s major hospitals indicated that close to 1/3 patients sought cancer care in India [Ref Wangai MW]. Millions of Americans go to other countries for medical services each year [ref CDC Yellow Book, Koons B]. 1/3 of Korean Americans in the Seattle area were reported to have gone to South Korea as medical tourists for services including colon cancer screening[ref Ko LK], and South Korean study showed that prevalence of GI cancers screened in Korean Americans was similar to that in native Koreans [ref Kim HS]. Knowledge and connection of immigrant cancer patients makes it more likely that they take advantage of lower-cost or culture-concordant care in other countries than someone born in the US [Ref Swami N].

The deaths of US citizens abroad are reported by US foreign missions in federal form DS-2060 (https://fam.state.gov/fam/07fam/07fam0270.html). The current version contains a cause-of-death field but no field for race/ethnicity. Therefore, these deaths are not counted in the US public health databases for mortality. Such missing data might lead to a potential under-reporting of 1/3 or above, based on the ASDRs of GI cancers for the Asian Indian group discussed earlier.

## DISCUSSION

### Advocacy for GI cancers as national priorities of health disparity

Gastrointestinal cancers contributed to more than a quarter of overall cancer deaths 2018-2022 in the U.S. [ref Kochanek KD]., yet these cancers that disproportionally affect minority race/ethnic groups remain to be to underfunded by NCI or non-profit organisations [ref Kamath SD], and often missing in national health disparity discussions ref Haghighat S]. CRC, stomach, bile duct, and liver cancers causes thousands of deaths in the younger populations (Supplement 1). Liver and stomach cancers are associated with preventable infectious agents, yet despite research and recommendations [Ref McCracken M, Huang RJ, Thompson CA, Volesky-Avellaneda KD], US has not yet implemented national guidelines to mitigate these risks or finding these cancers early as Japan successfully did [ref Kudo M]. We need to advocate for more awareness and research investment into these deadly GI cancers that often lack visible groups of survivor advocates.

### Missing Data as a Sign of Health Disparity

In this deep-dive and cross-examination of mortality data for GI cancers, the under-reporting of deaths among minority groups emerged as signs of health disparity. Under-reporting has affected our understanding of the US health equity landscape, making issues in vulnerable populations even less visible. In addition to the known issue of mis-assigned race/ethnic identity in death certificates, the reasons behind unreliable death numbers in non-White Hispanic and “More than one race” non-Hispanic group require further investigation. In this article, we identified the exclusion of US citizens’ death abroad as another issue of under-reporting, particularly for Asian groups with more recent immigrants whose birth countries have well-developed medical tourism industries. The Asian Indian group may be affected more than the Vietnamese group whose home country imposes more stringent visa requirements. The Hispanic population in US may also face similar under-reporting of cancer deaths and warrant further research, with disaggregated subgroup analysis (Mexican vs Cuban, for example).

### Global and Cross-Agency Efforts to Accurately Assess Cancer Health Disparity in Immigrants

With the 2003 version of death certificates implemented across the US since 2018, the public can now access nation-wide mortality data from CDC. Many recent publications have utilised the mortality data from WONDER@CDC for health disparity research [Ref Zhu DT, Wang CP, Mahmood S]. However, we can not assumed that the low death rates for some race/ethnic groups meant low cancer burden. We would like to call attention to seeing such unreasonably low numbers as a sign of disparity in reporting, and advocate for collaborations cross borders with researchers in immigrants’ birth countries to better understand the risk factors, epidemiological data, screening successes, and treatment options. We need more national investments into supporting immigrant cancer patients who face more barriers to access quality care in the US - language barriers, medical illiteracy, culture challenges, and financial burdens.

This issue of missing race/ethnic fields that is contributing to the under-reporting of deaths overseas can be resolved by updating the consulate report of American citizen death abroad (DS2060) to include the information similar to the 2003 version of death certificates. However, the data on DS2060 is collected by the Department of States, whereas the reporting is the responsibility of Center of Disease Control (belonging to the Department of Health and Human Services). Advocacy for additional resources will be needed to make sure US foreign missions have the capacities to incorporate such changes. Piloting in countries/regions where we see significant signs of under-reporting might help address the problem faster.

### Limitations of data quality and analysis

Race and ethnic attribution for different Asian groups in Single Race 15 grouping contained in US death certificates has not been assessed for accuracy as as those in Single Race 6 grouping [ref Fernandez Perez C], and the denominator we used to calculate ASDR of Asian groups was derived from Census instead of being provided by CDC. Disaggregation of death numbers in different sub-groups may be affected by WONDER’s data suppression rules to protect patients’ privacy - especially for cancers that were reported from several ICD codes. We used 5 years data (2018-2022) to overcome this issue. The total death numbers in table 2 was 0.2% higher than all the sub-groups combined for cancers in colon and rectum and 0.28% higher for stomach cancer. The impacts of suppression might be bigger for smaller populations and in smaller age groups, so we will need to wait for longer data collection period to run similar analysis for the HNOPI/NH population.

## Data Availability

All data produced in the present study are available upon reasonable request to the authors

## ACKNOWLEDGEMENT

We would like to thank NCI’s Office of Advocacy Relations, and Dr Meredith Shiels from NCI for discussions over potential data quality issues in the WONDER@CDC datasets.

